# No association between *FMR1* premutation and either ADHD or anxiety in 53,707 women undergoing genetic testing for family planning purposes

**DOI:** 10.1101/2024.10.06.24314952

**Authors:** Liraz Klausner, Shai Carmi, Shay Ben-Shachar, Noa Lev-El Halabi, Lina Basel-Salmon, Dana Brabbing Goldstein

## Abstract

**Background:** An *FMR1* full mutation, which causes Fragile X Syndrome, is defined as >200 repeats of the CGG motif in the gene’s 5’ untranslated region. A repeat count in the range 55-200 is considered an *FMR1* premutation (PM) and was previously associated with neuropsychiatric phenotypes. However, these associations did not always replicate and may be due to ascertainment bias. Here, we studied the association between PM and attention deficit hyperactivity disorder (ADHD) and anxiety using large population-based screening data.

**Methods:** We used data on women who underwent genetic screening in Rabin Medical Center in Israel for family planning purposes between 2001-2020. PM carriers were defined as subjects with 58-200 CGG repeats. We linked the genetic testing results to longitudinal electronic medical records (EMR) from Clalit Health Services. We defined ADHD and anxiety based on either a formal diagnosis or the purchase of relevant medications. As a positive control, we considered premature ovarian insufficiency (POI) and high follicle-stimulating hormone (FSH) levels before the age of 40. Our primary analysis used Cox regression with socioeconomic status, immigration, and age at testing as covariates.

**Results:** Our sample included 53,707 women: 464 PM carriers and 53,243 non-carriers. PM was associated with POI (hazard ratio (HR): 4.08, 95% confidence interval (CI): 2.16-7.72) and high FSH (HR: 3.43, 95% CI: 2.65-4.43). However, PM was not associated with either ADHD (HR 0.95; 95% CI: 0.51-1.77; 1331 events) or anxiety (HR 0.81; 95% CI: 0.47-1.39; 1814 events). The results were similar when the phenotype was defined based on medications and with logistic regression. Our study was sufficiently powered to detect HR about 2 or higher.

**Discussion:** We found no association between PM and either ADHD or anxiety. Our study is less prone to ascertainment bias towards affected families; however, the ascertained subjects are likely healthier than the population average. While our sample size is the largest to date, given the low frequency of PM carriers, small effects cannot be excluded.

## Introduction

Fragile X Syndrome (FXS) and other *FMR1* related disorders are caused by CGG trinucleotide repeat expansion in the 5’ untranslated region of the *FMR1* mRNA. An expansion to 200 or more copies is called *FMR1* full mutation. It causes hypermethylation and loss of function of *FMR1*, which in turn causes FXS — the most common cause of inherited intellectual disability and autism^1^. An expansion of 55-200 CGG repeats is considered an *FMR1* premutation (PM), with a female carrier rate previously estimated as 1 in 113-178^2,3^. It has been hypothesized that gain of function RNA toxicity is the main mechanism responsible for *FMR1* premutation associated conditions^4^.

While PM carriers do not have FXS, they are at high risk for several other conditions. The primary associated conditions are fragile X-associated tremor/ataxia syndrome (FXTAS) in both males and females (lifetime risk of 30-40% and 8-16%, respectively)^5^ and fragile X-associated primary ovarian insufficiency (FXPOI) in about 20% of females^6^. Over the last two decades, a broad spectrum of other medical conditions has been described in PM carriers. Several studies observed that female PM carriers are at increased risk for neuropsychiatric disorders (FXAND), including anxiety disorders, depression, obsessive–compulsive behavior, attention deficit hyperactivity disorder (ADHD) inattentive type, and autistic spectrum phenotypes^7–14^ It has been suggested that those symptoms are more apparent or frequent in older carriers, in patients with FXTAS, and in mid-size CGG repeat number^15–17^. However, other studies did not replicate these associations^18–21^

Whether or not neuropsychiatric conditions are associated with PM is the subject on an ongoing debate. Some argue that the apparent associations result from ascertainment bias, given that PM carriers are often identified only after a family member has been diagnosed with FXS. PM carriers ascertained this way may carry a higher number of repeats, have predisposing genetic modifiers, or suffer from exposure to the challenges of caring for someone with FXS^8,10^.

Here, we contribute to this debate by studying data from population-wide screening, which is less prone to ascertainment bias towards affected families. Specifically, we study neuropsychiatric conditions, as identified by electronic medical records (EMR), in >50,000 women who underwent voluntary free-of-charge screening for autosomal recessive disorders and two X-linked disorders including Fragile X syndrome^22^. We find no association between PM and neuropsychiatric conditions. However, given the low frequency of PM, our study was only powered to detect large effects.

## Methods

### Ethics

The study was approved by the Institutional Review Board of Rabin Medical Center, approval number 0197-20.

### Study population

Our study population consisted of 54,447 women in their reproductive years who underwent genetic screening offered at no cost by the Israel Ministry of Health for family planning purposes. The test was conducted by the Genetic institute at Rabin Medical Center in Israel for members of Clalit Health Services between the end of 2001 and the end of 2020. The screening program is offered to all women in their reproductive years regardless of family history, thereby substantially reducing confounding due to ascertainment bias.

### Testing method

The *FMR1* test was performed using a Southern Blot analysis until 2007, a TR-PCR analysis during the years 2007-2010, and using an AmplideX PCR/CE FMR1 Kit by Assuragen from 2010 and on. The kit provides a PCR-only approach based on Triplet Repeat Primed PCR (TP-PCR) design to reliably amplify and detect all alleles, including full mutations.

### Defining the exposure

According to the recommendation of the Israeli Society of Medical Genetics and the policy of the Israel Ministry of Health, PM is defined as CGG repeat number in the range 58-200 (https://www.gov.il/he/departments/general/disease-genetic-fxs) in one of the two alleles. In case the experimental result returned a range for the repeat count, we used the upper limit of the range. As only 145 carriers had repeat count >70, we did not perform subgroup analyses on those with high repeat count.

### Exclusion criteria

Since our study focuses on premutation carriers, we removed 31 females who had more than 200 CGG repeats. To minimize non-screening-related referrals, which may lead to spurious associations (Figure S1), we excluded 616 females of age 18 or younger (4/616 were carriers) and 80 women over 50 (19/80 were carriers) at the time of genetic testing. We further excluded six women who had no data on dates of membership at Clalit and one woman who had an unknown number of repeats.

### Electronic medical records

We extracted anonymous EMR from Clalit Health Services using Clalit’s data sharing platform powered by MDclone (https://www.mdclone.com). The EMR was up to date up to September 7^th^, 2022. We searched the EMR for diagnoses related to neurocognitive and neuropsychological conditions and diagnoses related to women’s health and infertility. We show the overall number of women and the number of carrier women with each diagnosis in Table S1. We also extracted from the EMR demographic variables, lab tests, BMI, and purchased medications for ADHD, depression and anxiety, psychosis, epilepsy, and chronic pain.

### Defining the outcomes

We focused in this work on two relatively common neuropsychiatric conditions: ADHD and anxiety. We also considered premature ovarian insufficiency (POI), a known complication in PM carriers^23^, to serve as a positive control.

We defined the ADHD and anxiety phenotypes in two ways. (1) Based on a formal diagnosis; and (2) based on either having a formal diagnosis or the use of corresponding medications, specifically, purchasing any disorder-related drug at least three times. The relevant variables are listed in Table S2. Women were defined as having POI either based on a formal diagnosis or based on having follicle-stimulating hormone (FSH) levels over 15 IU/L under the age of 40.

In the statistical analysis, the time of the disease event was set as the time of diagnosis, if it exists, or otherwise the date of first drug purchase. For POI, it was set as the time of the first high FSH test.

### Statistical analysis

Events are censored outside the study period and in periods when subjects were not members at Clalit. We therefore used Cox regression to identify associations between being a carrier and having an increased hazard for the disease. As a robustness test, we used logistic regression with age as a covariate. In both analyses, we also included as covariates the socioeconomic score (one variable for each non-reference category + whether missing), whether the woman has immigrated, and the age (in years) at the time of genetic testing.

We accounted for membership at Clalit using the following methods. Among our subjects, 33,606 had a single continuous membership period at Clalit. Among other women, most membership gaps were due to military service (Figure S2). During service, which is compulsory at age 18 for a large proportion of Israeli women, soldiers no longer receive health services from their provider. We defined gaps due to military service as membership breaks lasting less than five years and overlapping with ages 18-21, identifying 16,307 women as having such gaps. In the regression analysis, these periods were not considered as gaps. The remaining women (3,806) had two separate membership periods. For these women, we considered the membership periods individually. Events that were not dated were not considered in the regression.

### Power analyses

While our overall sample size is very large, only 1% of the women are PM carriers. Therefore, we performed a power analysis to determine the effect sizes that are discoverable using our cohort. Denote by log Δ the log hazard ratio, p the proportion of carriers, and *z_l-α/2_* and *z_l-β_* the quantiles of a standard normal distribution corresponding to type I and type II error rates α and β, respectively. Using a formula from^24^, the total number of events required for achieving the desired power is *N_e_ = (Z_1 − α/2_ + Z_1 − β_^2^ [p(1 − p)(logΔ)^2^]^−1^*. Solving it for the hazard ratio, we obtain Δ = exp[(Z_1 − α/2_ + Z_1 − β)_/√.N_e_p(1 − p) We use α = 0.05 and p = 0.01 and vary N_e_ based on the number of observed events. This calculation corresponds to a best-case scenario, where none of the other covariates has any predictive power.

## Results

The study population included 54,447 women who underwent genetic screening at Rabin Medical Center, Israel, between 20 1-2020. After applying a number of exclusion criteria (particularly, excluding women 18 years old or younger), we were left with 53,707 women: 464 PM carriers and 53,243 non-carriers (carrier rate 0.85%). The median repeat number of PM carriers was 64 (see Figure 1 for the distribution of the PM repeat number). Table 1 reports summary statistics for the main characteristics of carriers and non-carriers. Carriers were slightly older (both at data extraction and at genetic testing) and had a slightly higher socioeconomic status. The immigration status and year of genetic testing were similar.

**Figure 1:**
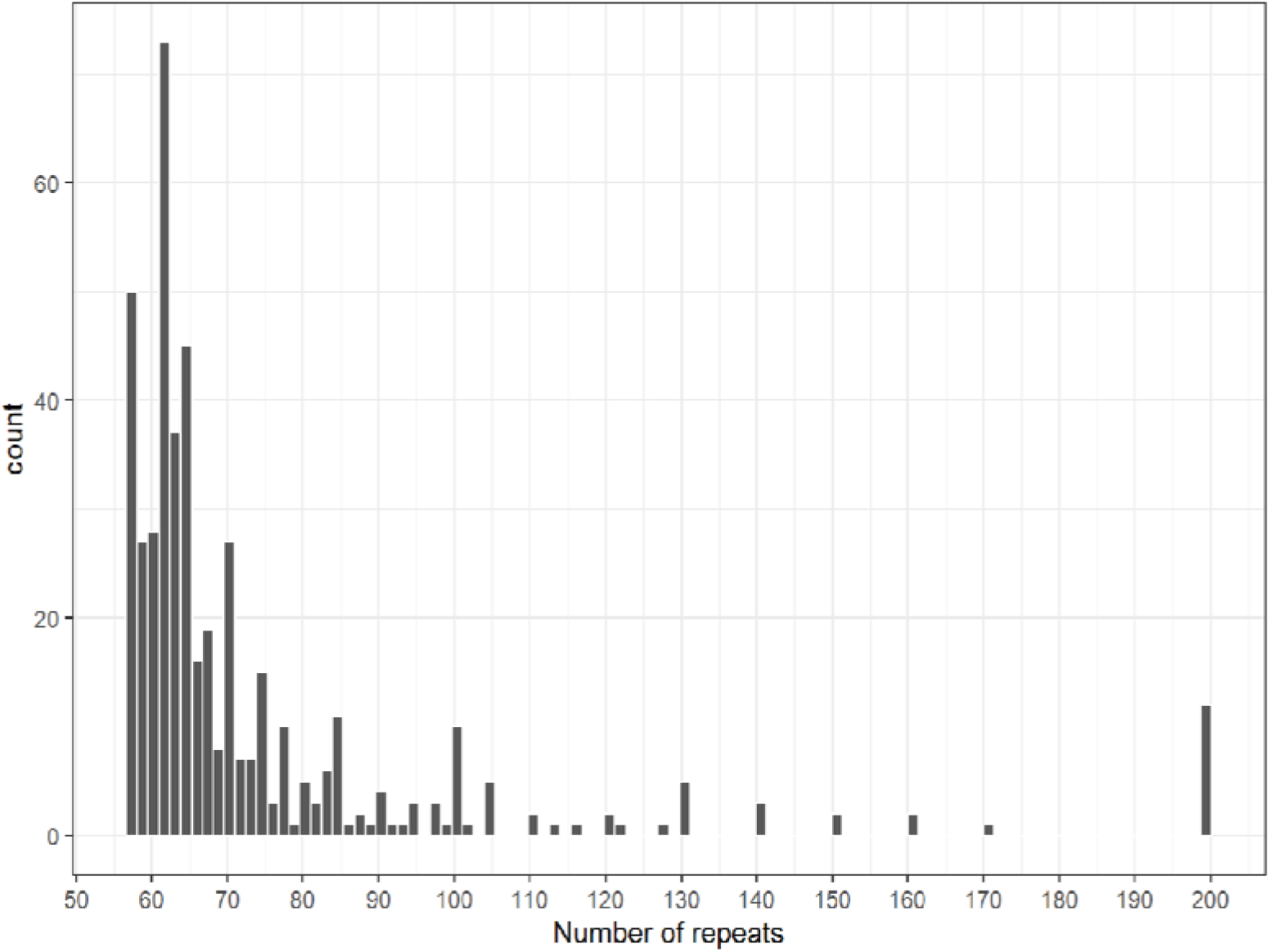
A histogram of the repeat number of PM carriers.

**Table 1:** Characteristics of our study population, broken down by *FMR1* repeat PM carrier status. The P-values are based on the Mann-Whitney U test for the continuous variables and the chi-squared test for the categorical variables.

| Parameter | Carriers | Non-carriers | P-value |
| --- | --- | --- | --- |
| Sample size (N) | 464 | 53243 |  |
| Age (median, IQR) | 40 (35-45) | 38 (34-43) | 3.2e-9 |
| Socioeconomic status (%) |  |  |  |
| Very Low | 3% | 6% | 1.9e-6 |
| Low | 12% | 13% |  |
| Medium | 25% | 29% |  |
| High | 39% | 35% |  |
| Very High | 16% | 10% |  |
| Missing | 5% | 7% |  |
| Immigrated (%) | 8% | 11% | 0.078 |
| Genetic test year (median, IQR) | 2014 (2010-2017) | 2014 (2011-2016) | 0.009 |
| Age at genetic test (median, IQR) | 31 (28-35) | 29 (26-33) | <1e-16 |

We compared the prevalence of ADHD, anxiety, and POI between carriers and non-carriers (Table 2). As expected, POI was at least 3 times more prevalent in carriers. In contrast, the prevalence of ADHD was about the same between the two groups, and the prevalence of anxiety was higher in non-carriers.

**Table 2:**
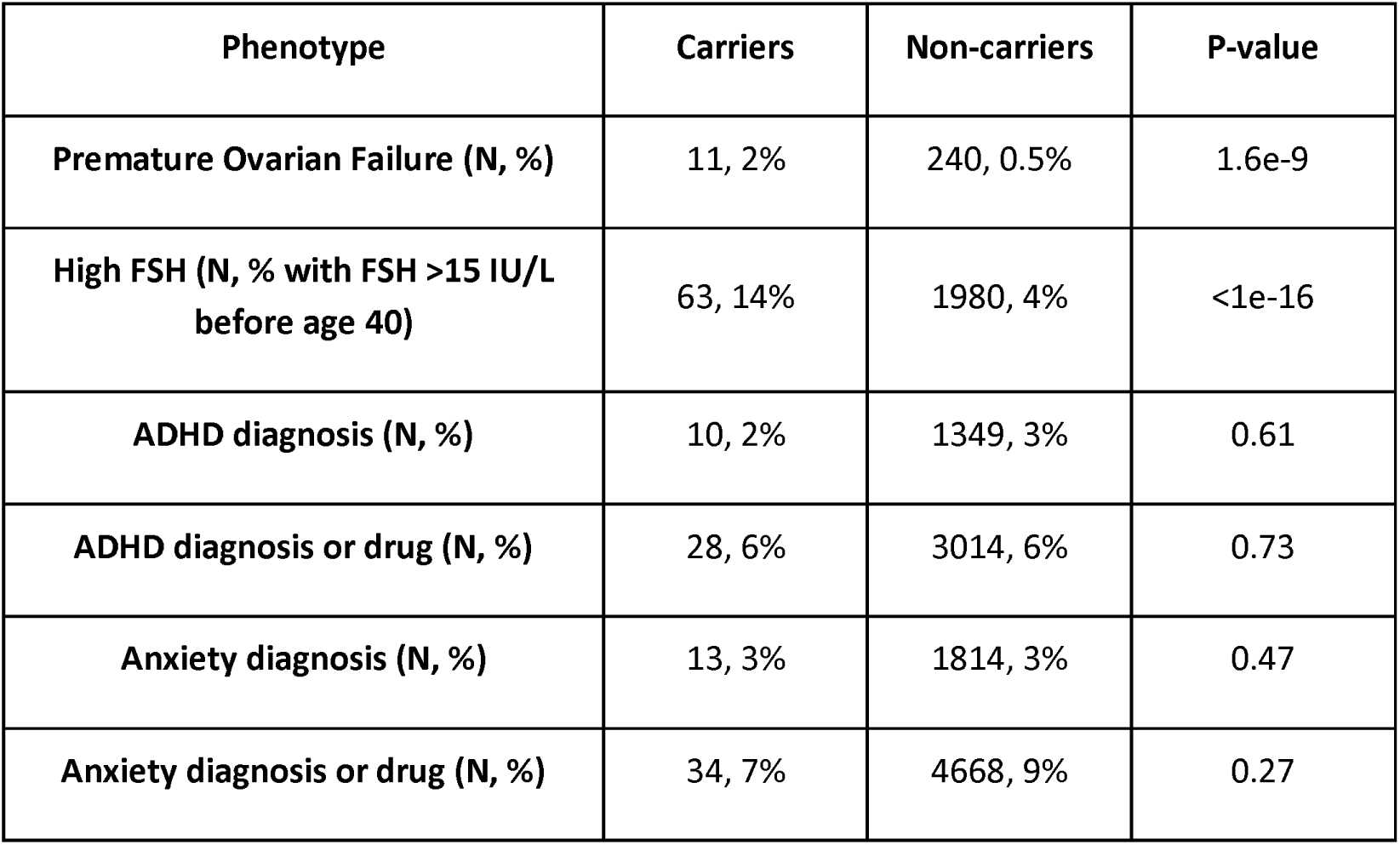
The prevalence of our target outcomes in carriers and non-carriers of *FMR1* PM. The P-values are based on chi-squared test. The event counts include a small number that were not dated and were thus not included in the Cox regressions below.

To account for covariates and for any differences in age distribution between carriers and non-carriers, we used Cox regression. Events were placed at time of diagnosis, or, if there was no diagnosis, at the first drug purchase (Methods). For premature ovarian failure (247 events across both carriers and non-carriers), being a carrier had a hazard ratio (HR) of 4.08 (95% CI: 2.16-7.72; Table 3; Figure S3). None of the covariates had a significant effect on the phenotype. The HR for high FSH was 3.43 (95% CI: 2.65-4.43; Table 3; 2039 events; Figure S4).

**Table 3:** Cox regression for premature ovarian failure. The estimates show the hazard ratios (HR). The reference socioeconomic level was medium.

|  | Premature ovarian failure diagnosis |  |  | High FSH (> 15 IU/L) |  |  |
| --- | --- | --- | --- | --- | --- | --- |
|  | HR | 95% CI | P-value | HR | 95% CI | P-value |
| Carrier | 4.08 | 2.16-7.72 | 1.5e-5 | 3.43 | 2.65-4.43 | <1e-16 |
| Socioeconomic - Very low | 1.29 | 0.7-2.39 | 0.41 | 1.02 | 0.82-1.28 | 0.84 |
| Socioeconomic - Low | 1.35 | 0.89-2.06 | 0.16 | 1.21 | 1.04-1.4 | 0.01 |
| Socioeconomic - High | 0.98 | 0.71-1.34 | 0.88 | 0.98 | 0.88-1.19 | 0.73 |
| Socioeconomic - Very high | 1.24 | 0.83-1.87 | 0.3 | 1.02 | 0.88-1.19 | 0.77 |
| **Socioeconomic - Missing** | 0.82 | 0.43-1.55 | 0.54 | 0.87 | 0.7-1.07 | 0.19 |
| **Immigrated** | 1.19 | 0.83-1.72 | 0.34 | 0.78 | 0.67-0.91 | 1.33e-3 |
| **Test age** | 1.01 | 0.99-1.04 | 0.26 | 1 | 0.99-1.01 | 0.5 |

In contrast to ovarian failure, we found no association between PM carrier status and ADHD or anxiety. In Cox regression for ADHD (Table 4), being a carrier had a statistically insignificant hazard ratio, either when ADHD was defined based on an EMR diagnosis (HR 0.95; 95% CI: 0.51-1.77; 1331 events; Figure S5) or when it was defined based on either diagnosis or medications (HR 1.08; 95% CI: 0.75-1.56; 3023 events; Figure S6). Here, multiple covariates had a significant effect. In Cox regression for anxiety (Table 5), being a carrier similarly had statistically insignificant hazard ratios either based on diagnosis (HR 0.81; 95% CI: 0.47-1.39; 1814 events; Figure S7) or based on diagnosis or drug (HR 0.74; 95% CI: 0.53-1.04; 4680 events; Figure S8). The results were similar with logistic regression (Tables S3 and S4), showing no significant effect of PM carrier status on either ADHD or anxiety.

**Table 4:** Cox regression for ADHD. The estimates show the hazard ratios. The reference socioeconomic level was medium.

|  | **ADHD diagnosis** |  |  | **ADHD diagnosis or medications** |  |  |
| --- | --- | --- | --- | --- | --- | --- |
|  | **HR** | **95% CI** | **P-value** | **HR** | **95% CI** | **P-value** |
| **Carrier** | 0.95 | 0.51-1.77 | 0.88 | 1.08 | 0.75-1.56 | 0.68 |
| **Socioeconomic - Very low** | 0.08 | 0.04-0.15 | 4.68e-15 | 0.06 | 0.03-0.1 | <1e-16 |
| **Socioeconomic - Low** | 0.71 | 0.59-0.86 | 2.87e-4 | 0.66 | 0.57-0.75 | 7.49e-10 |
| **Socioeconomic - High** | 1.22 | 1.08-1.39 | 2.12e-3 | 1.34 | 1.23-1.46 | 2.84e-11 |
| **Socioeconomic - Very high** | 1.23 | 1.02-1.49 | 0.03 | 1.44 | 1.28-1.62 | 1.71e-09 |
| **Socioeconomic - missing** | 0.51 | 0.39-0.67 | 1.34e-6 | 0.56 | 0.47-0.68 | 1.86e-09 |
| **Immigrated** | 0.67 | 0.54-0.84 | 4.14e-4 | 0.76 | 0.66-0.87 | 6.25e-05 |
| **Test age** | 0.95 | 0.94-0.97 | 1.54e-11 | 0.98 | 0.97-0.99 | 1.32e-05 |

**Table 5:** Cox regression for anxiety. The estimates show the hazard ratios. The reference socioeconomic level was medium.

|  | Anxiety diagnosis |  |  | Anxiety diagnosis or medications |  |  |
| --- | --- | --- | --- | --- | --- | --- |
|  | HR | 95% CI | P-value | HR | 95% CI | P-value |
| **Carrier** | 0.81 | 0.47-1.39 | 0.45 | 0.74 | 0.53-1.04 | 0.08 |
| **Socioeconomic - Very low** | 0.67 | 0.52-0.88 | 3.23e-3 | 0.63 | 0.53-0.74 | 8.57e-08 |
| **Socioeconomic - Low** | 0.95 | 0.82-1.12 | 0.56 | 0.97 | 0.88-1.08 | 0.62 |
| **Socioeconomic - High** | 0.86 | 0.77-0.96 | 6.62e-3 | 0.99 | 0.92-1.06 | 0.75 |
| **Socioeconomic - Very high** | 0.68 | 0.57-0.81 | 1.23e-05 | 0.97 | 0.88-1.08 | 0.6 |
| **Socioeconomic - missing** | 0.99 | 0.82-1.21 | 0.97 | 0.95 | 0.83-1.08 | 0.41 |
| **Immigrated** | 1.07 | 0.93-1.23 | 0.33 | 1.19 | 1.09-1.29 | 8.69e-05 |
| **Test age** | 0.94 | 0.93-0.95 | <1e-16 | 1.03 | 1.03-1.04 | <1e-16 |

The null results we observed may be due to low power. Our power analysis (Methods) suggests that with the ≈1300-1800 events of ADHD and anxiety based on diagnosis alone, we require a hazard ratio of at least 1.94-2.18 to have 80% power. When using either diagnosis or drugs, with ≈3000-4700 events, we need a hazard ratio of 1.51-1.67. Therefore, while our results argue strongly against hazard ratios >2, we cannot exclude smaller effects.

## Discussion

Whether *FMR1* premutation is associated with neuropsychiatric disorders is still under debate, where some studies demonstrated correlation^8–10^ but others did not^18,19^. Here, we leveraged data from over 50,000 women from Israel to add to the debate, finding no association between PM and either ADHD or anxiety. To the best of our knowledge, ours is the largest population-based study to examine these associations. Our study should have had sufficient power to detect associations with HR about 2 or higher (as previously observed for ADHD and anxiety^21^), and we demonstrated its ability to detect the correlation between PM and premature ovarian insufficiency. A limitation of our study is the focus only on ADHD and anxiety, given the rarity of other conditions among carriers, which means that our results may not necessarily generalize to all neuropsychiatric disorders.

In contrast to most previous studies, our cohort was population-based, namely women who took a genetic test offered to the general population in the absence of medical indications. This study design is less prone to ascertainment bias towards families with at least one affected member or individuals with preexisting conditions or symptoms, which may explain the previously observed associations. However, our “genotype-first” approach may generate ascertainment bias towards subjects healthier than the average^25^ which was shown to lead, across various disorders, to lower estimates of penetrance compared to clinical cohorts^26,27^. In particular, most of our subjects were genetically tested for family planning purposes, implying they were healthy enough to start a family and were aware of the risk of genetic disorders. While the uptake of preconception genetic screening is very high in Israel, particularly in the Jewish sector^28^, our sample is still not a random sample from the population. On the other hand, our study was able to detect the association of PM with POI.

There are additional reasons that may explain the difference between our study and previous efforts. First, the group of PM carriers is heterogeneous, particularly with respect to repeat count. For example, the mean repeat number in PM carriers from cohorts based on referral or cascade testing ^11,12^ was 92-97, substantially higher than in our cohort (74). In the population-based study of Movaghar et al, which found an association between PM and anxiety, the average repeat number was 67^8^, similar to ours. However, that study excluded individuals with 7-23 and 41-54 repeats, which are expected to contribute to the asymptomatic group. The repeat count distribution may affect conclusions, as it is associated with symptoms. For example, Schmitt et al identified three groups of PM carriers with differences in repeat count, neuropsychiatric conditions, and executive function^19^, and Gabis et al found an increase in learning difficulties and daily function with increasing repeat count^14^.

Second, neuropsychiatric symptoms previously associated with PM are often mild and may have not been recorded in our EMR. Indeed, as many symptoms do not fulfill the DSM definition of a neuropsychiatric disorder, they were previously suggested to be called “Fragile X Associated Conditions”^29^. For example, one study of female PM carriers identified mild learning and attention difficulties that impacted daily functioning^14^. These subtle difficulties may go unnoticed without targeted testing or specific awareness, and therefore may not be documented in EMR.

The results of our study have implications for genetic counselling of patients who have discovered an *FMR1* PM during preconception or prenatal screening. Along with previous work^18,20^, our results provide some assurance regarding the risk of neuropsychiatric disorders for the fetus or a future child. However, it is important to consider that (i) several previous studies did find an association; (ii) studies of PM and neuropsychiatric conditions have varied in methodology, terminology, and severity of conditions^10^; and (iii) many additional factors may contribute to these manifestations^13^. Therefore, further research is needed, particularly prospective studies of carrier fetuses and newborns, before definitive conclusions are drawn for genetic counseling and patient management.

## Data Availability

The electronic medical records analyzed as part of this study are unavailable to external researchers.

## Acknowledgements

We thank Hila Fridman for discussions. L.K. was supported by a United States-Israel Binational Science Foundation grant no. 2017024 to S.C.

## Supplementary figures

**Figure S1:**
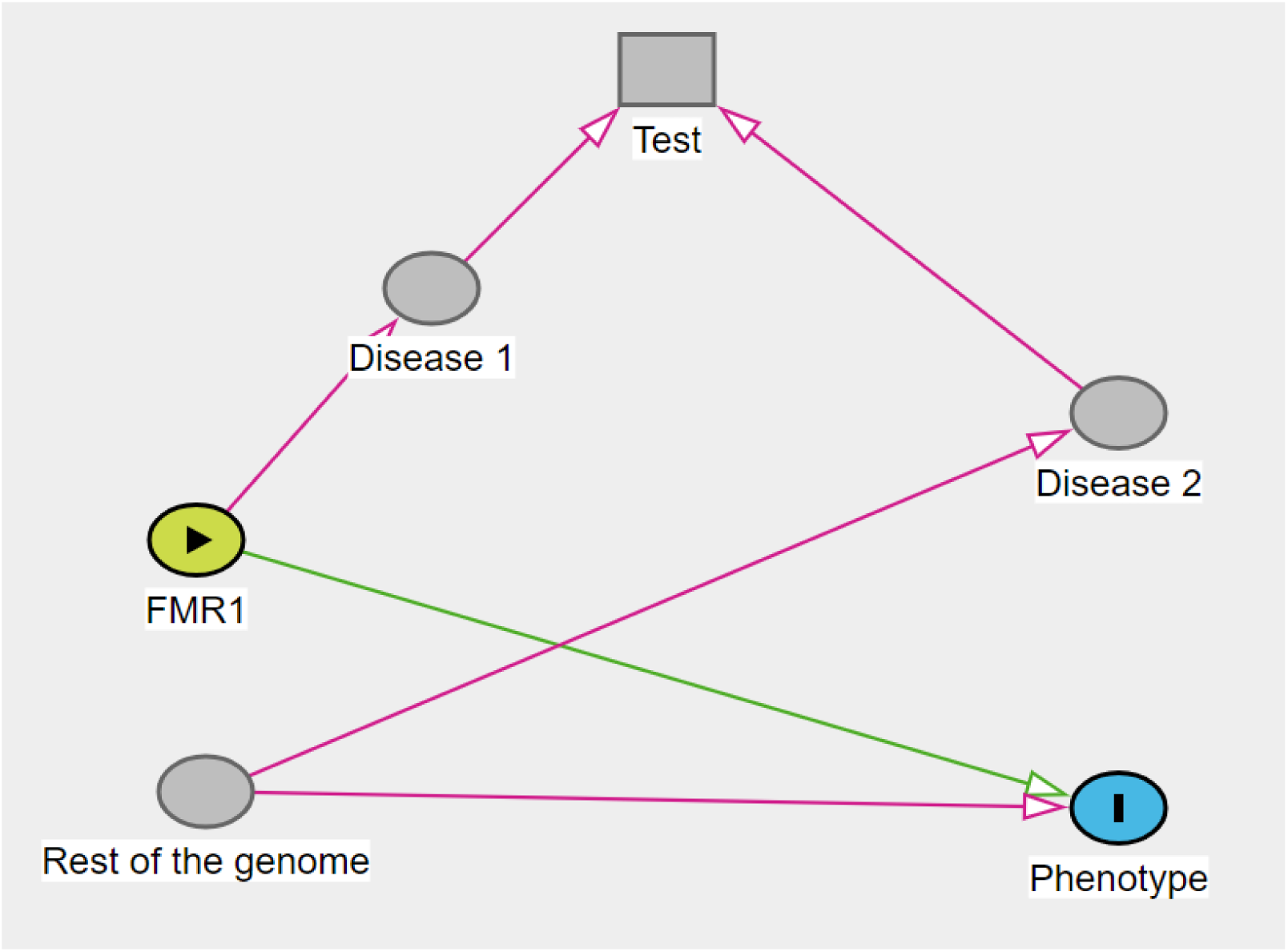
A directed acyclic graph demonstrating the possibility of a spurious association between the FMR1 carrier status and the phenotype when genetic testing is performed on indication. The graph was visualized and analyzed using DAGitty (https://www.dagitty.net/dags.html). FMR1 is the exposure (yellow). The target phenotype is the outcome (light blue). We are interested in the causal relationship between FMR1 PM and the target phenotype. However, PM may also cause other conditions, which may affect the likelihood of genetic testing (square). If there is another genetic variant affecting both the target phenotype and another condition leading to genetic testing, the test becomes a collider, opening a non-causal path between FMR1 and the target phenotype. The same holds in case the target phenotype itself affects the likelihood of a genetic test.

**Figure S2:**
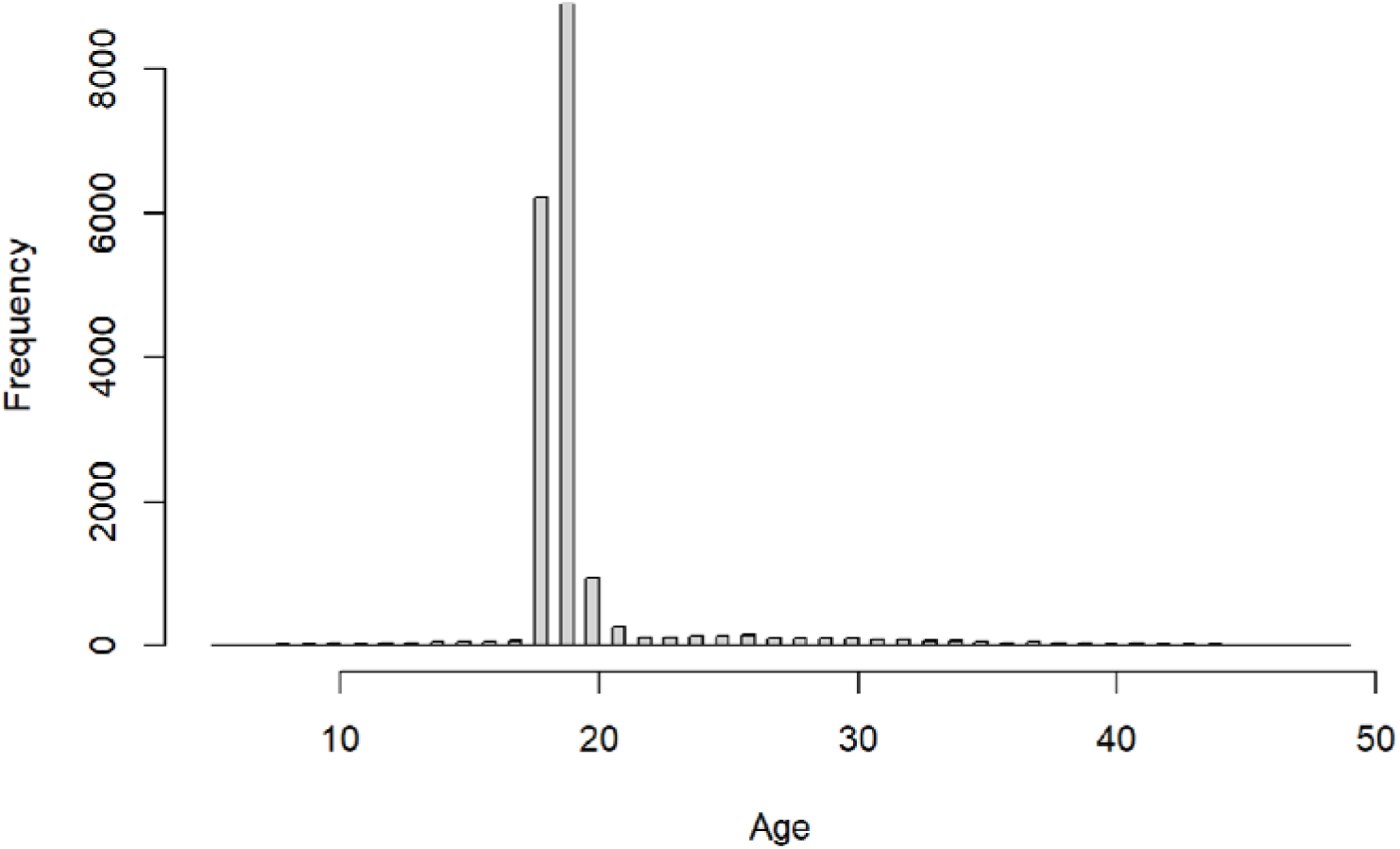
The distribution of ages when a membership gap lasting five years or less has started. Most membership breaks start between the ages of 18 and 21, in agreement with attributing these gaps to a mandatory military service.

**Figure S3:**
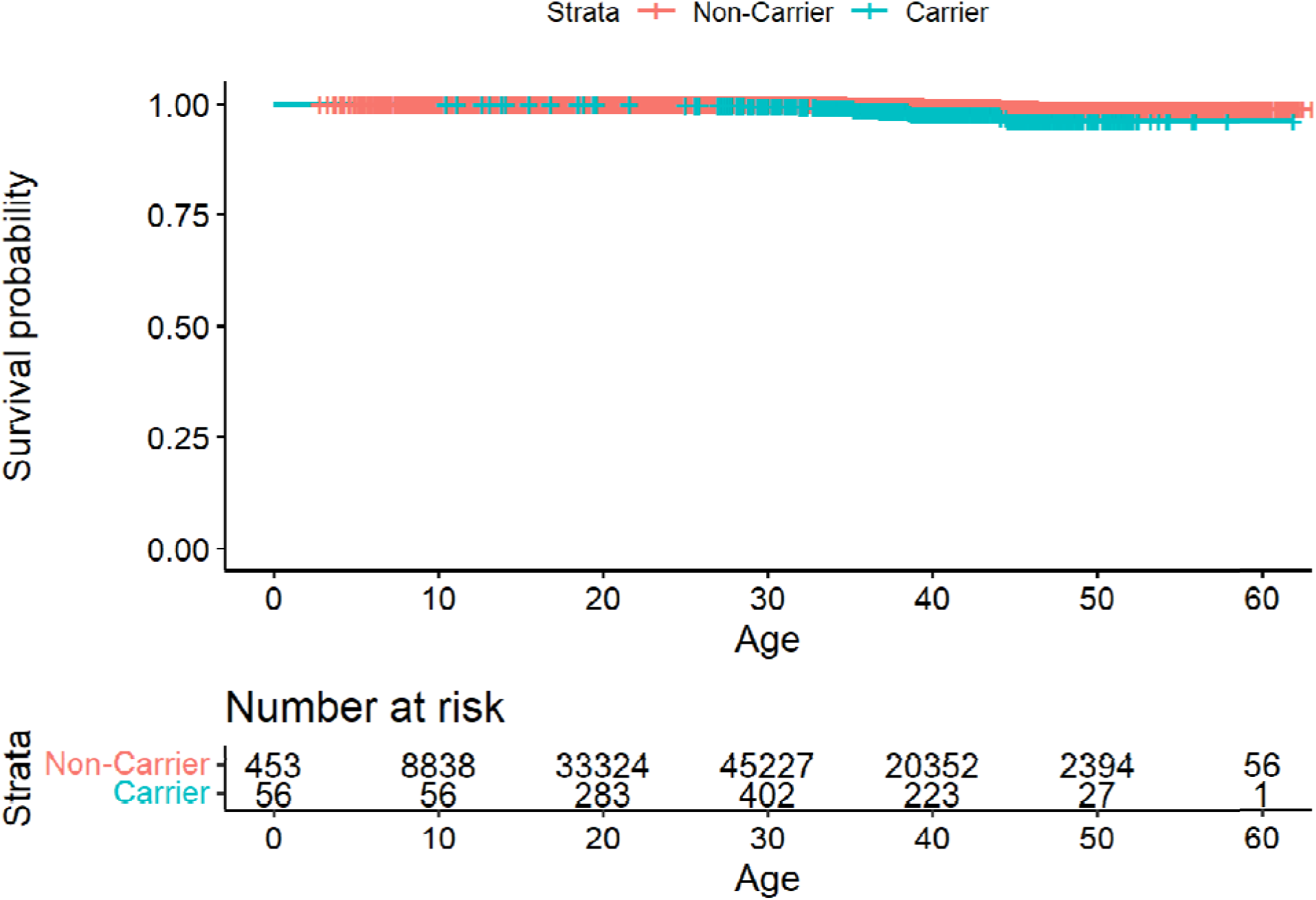
A Kaplan-Meier survival curve for premature ovarian failure.

**Figure S4:**
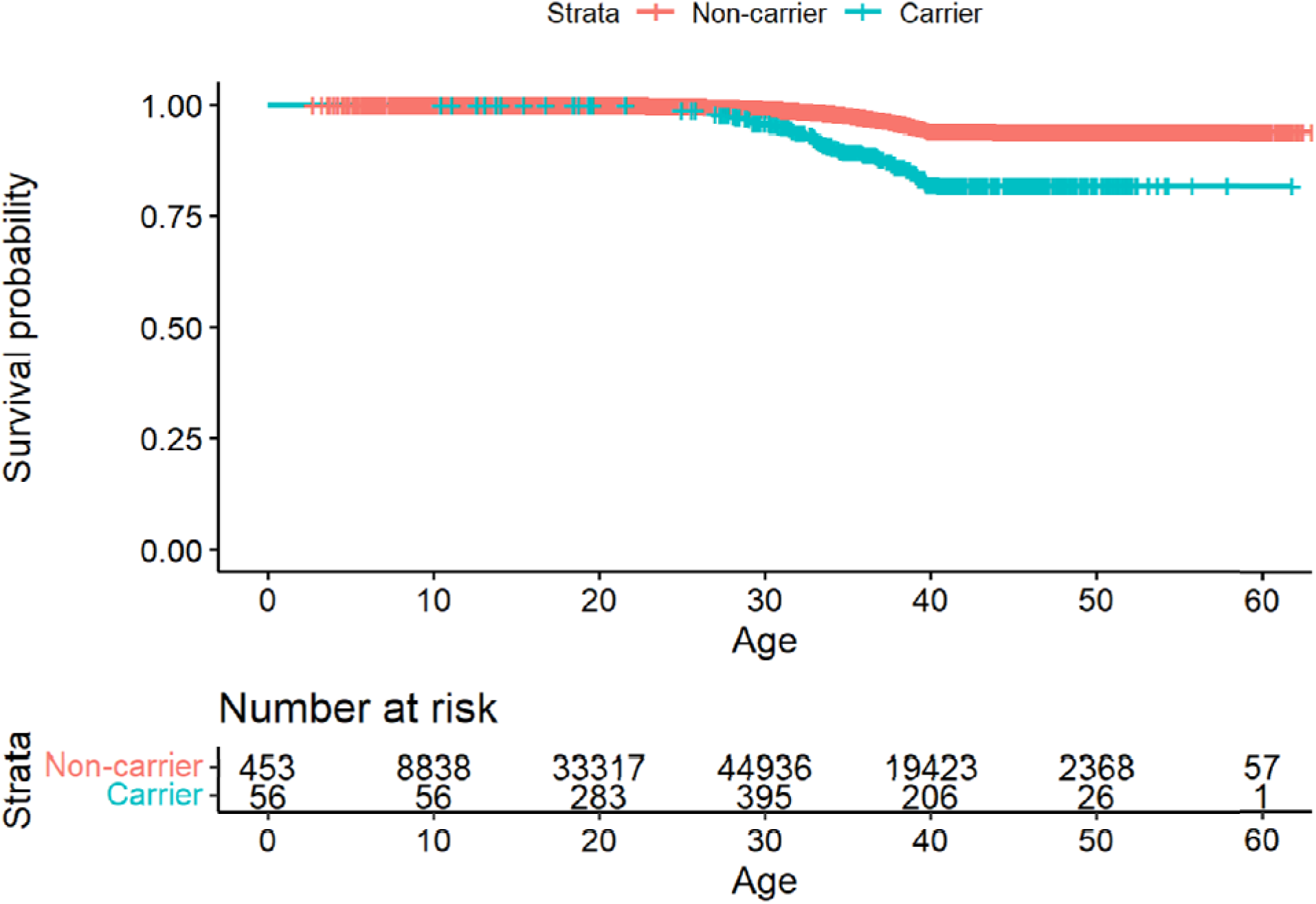
A Kaplan-Meier survival curve for FSH. Note that no events can be recorded past age 40, given that an event is defined as “having follicle-stimulating hormone (FSH) levels over 15 IU/L under the age of 40”.

**Figure S5:**
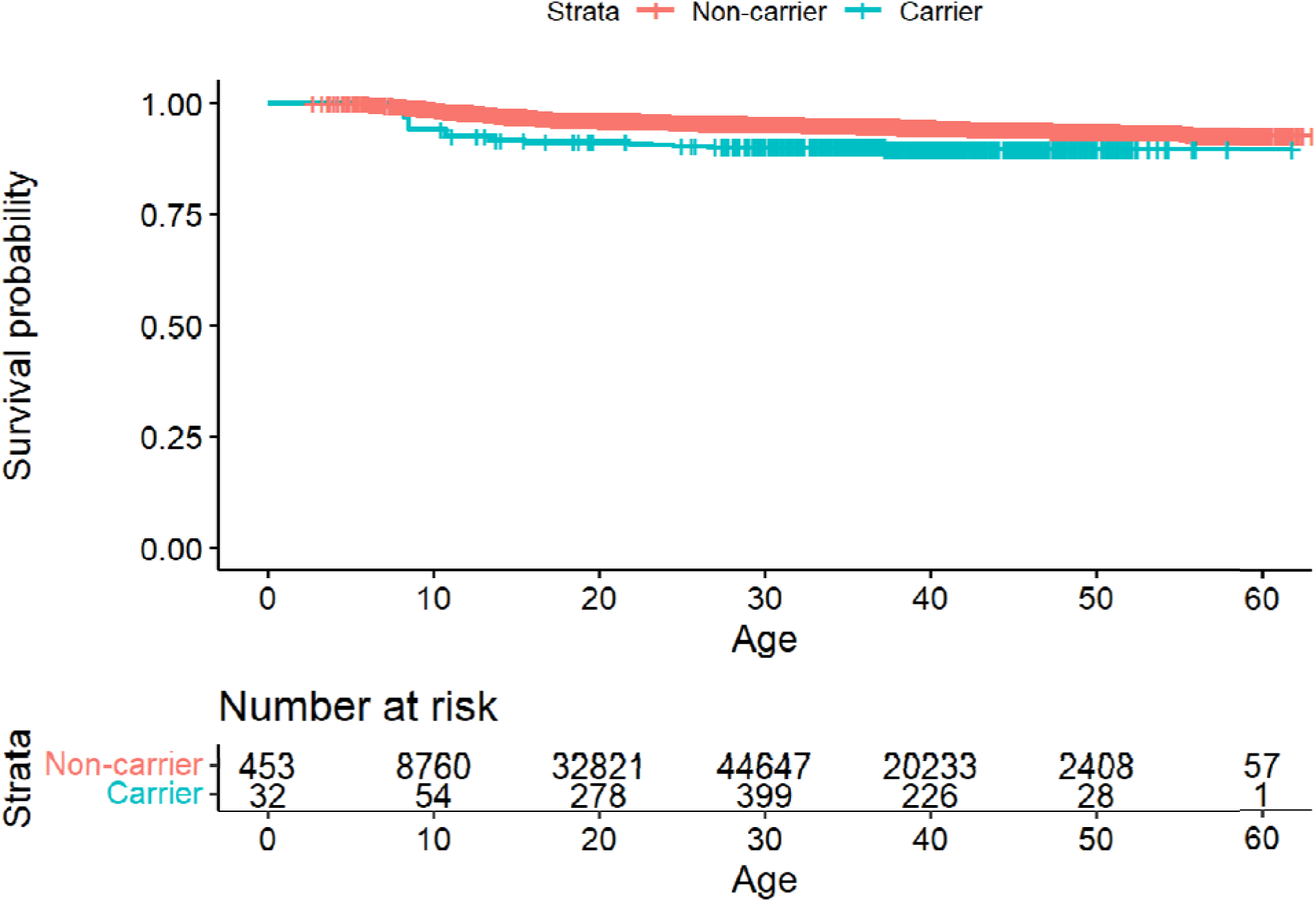
A Kaplan-Meier survival curve for ADHD diagnosis.

**Figure S6:**
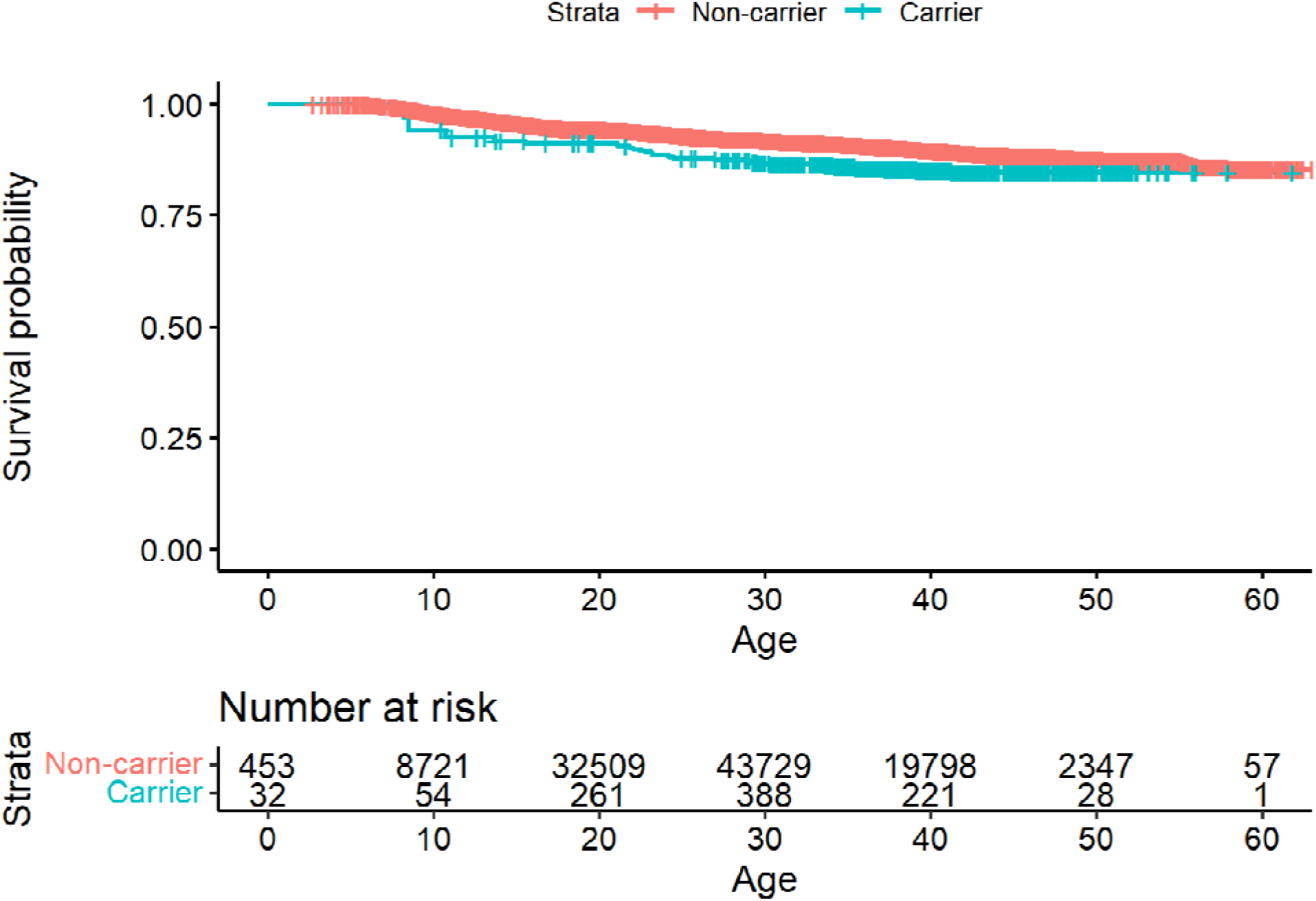
A Kaplan-Meier survival curve for ADHD based on diagnosis or medications.

**Figure S7:**
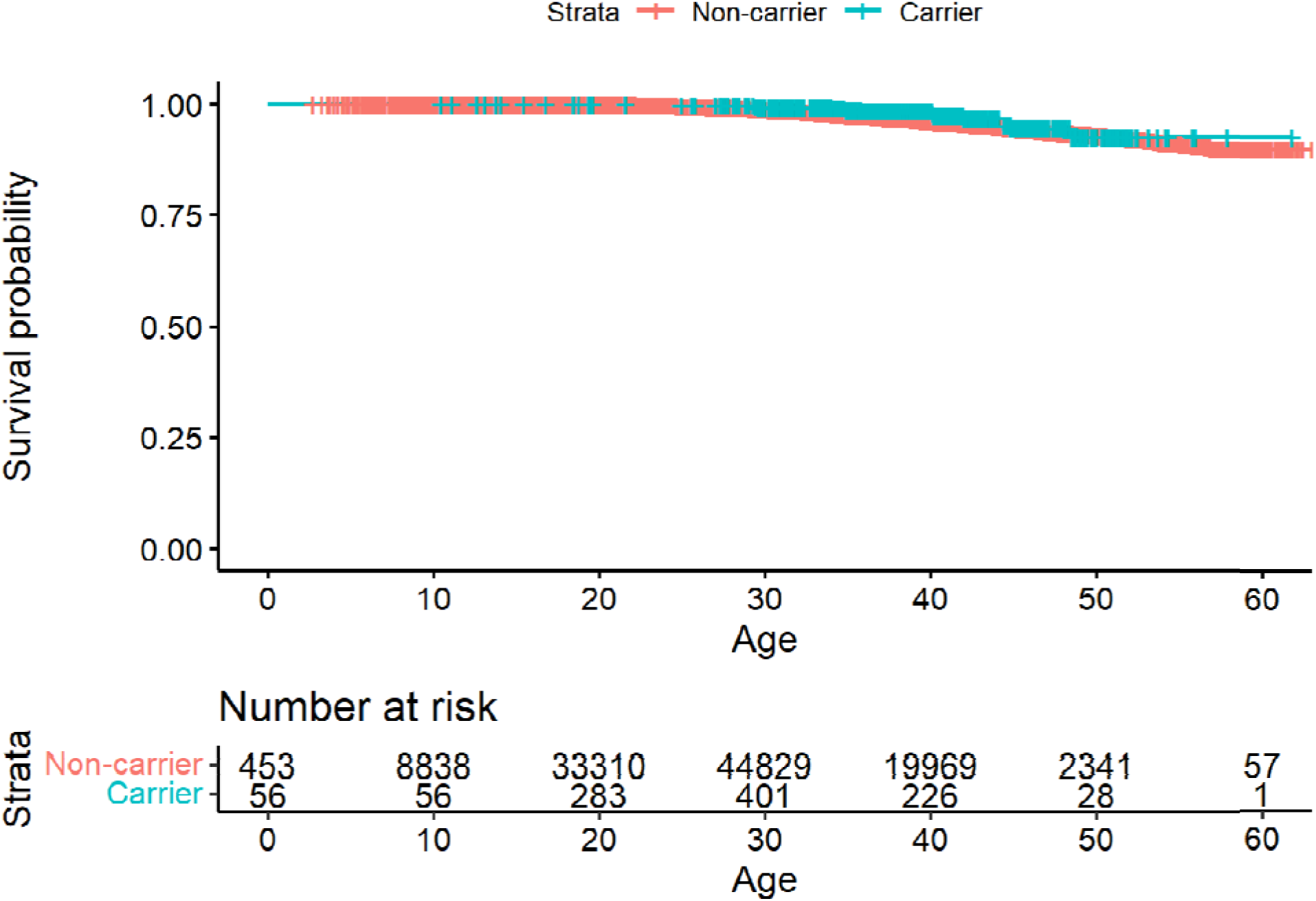
A Kaplan-Meier survival curve for anxiety diagnosis.

**Figure S8:**
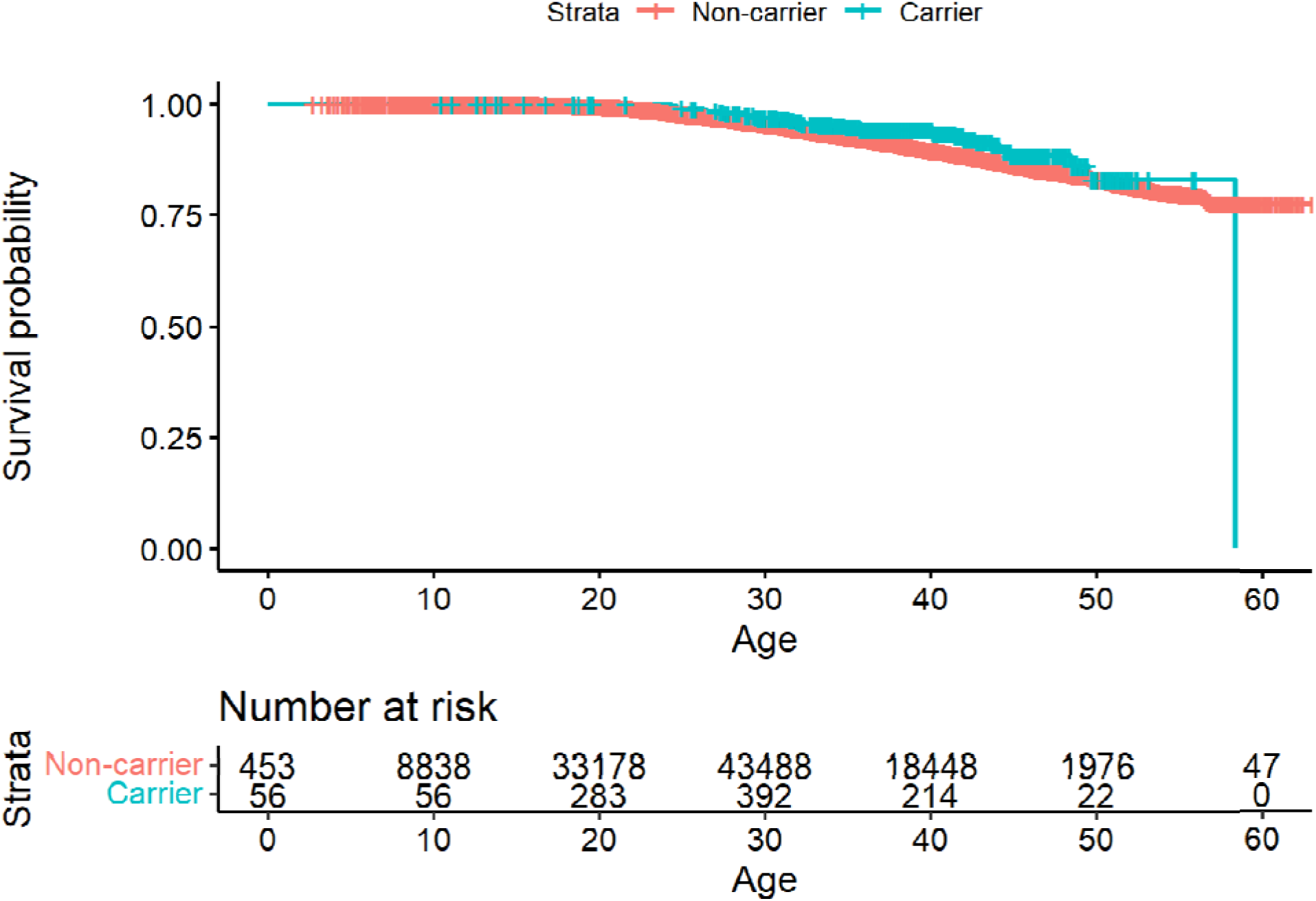
A Kaplan-Meier survival curve for anxiety based on diagnosis or medications.

## Supplementary tables

**Table S1.** The number of cases for each diagnosis. (See Excel file)

**Table S2.** A list of all variables available for this study, including demographic information, diagnoses, and medications. (See Excel file)

**Table S3.** Logistic regression for ADHD.

|  | ADHD diagnosis |  |  | ADHD diagnosis or medications |  |  |
| --- | --- | --- | --- | --- | --- | --- |
|  | log(OR) | SD | P-value | log(OR) | SD | P-value |
| **Intercept** | -1.34 | 0.18 | 2.32e-14 | -1.72 | 0.12 | <1e-16 |
| **Carrier** | -0.09 | 0.32 | 0.78 | 0.05 | 0.2 | 0.81 |
| **Socioeconomic - Very low** | -2.53 | 0.32 | 3.46e-15 | -2.9 | 0.28 | <1e-16 |
| **Socioeconomic - Low** | -0.29 | 0.09 | 2.37e-3 | -0.39 | 0.07 | 1.98e-8 |
| **Socioeconomic - High** | 0.16 | 0.07 | 0.01 | 0.28 | 0.05 | 4.07e-10 |
| **Socioeconomic - Very high** | 0.16 | 0.1 | 0.09 | 0.35 | 0.06 | 1.98e-8 |
| **Socioeconomic - missing** | -0.67 | 0.14 | 1.4e-6 | -0.6 | 0.1 | 6.07e-10 |
| **Immigrated** | -0.6 | 0.11 | 9.13e-8 | -0.45 | 0.07 | 1.12e-10 |
| **Test age** | -0.08 | 0.01 | <1e-16 | -0.04 | 3.9e-3 | <1e-16 |

**Table S4.**
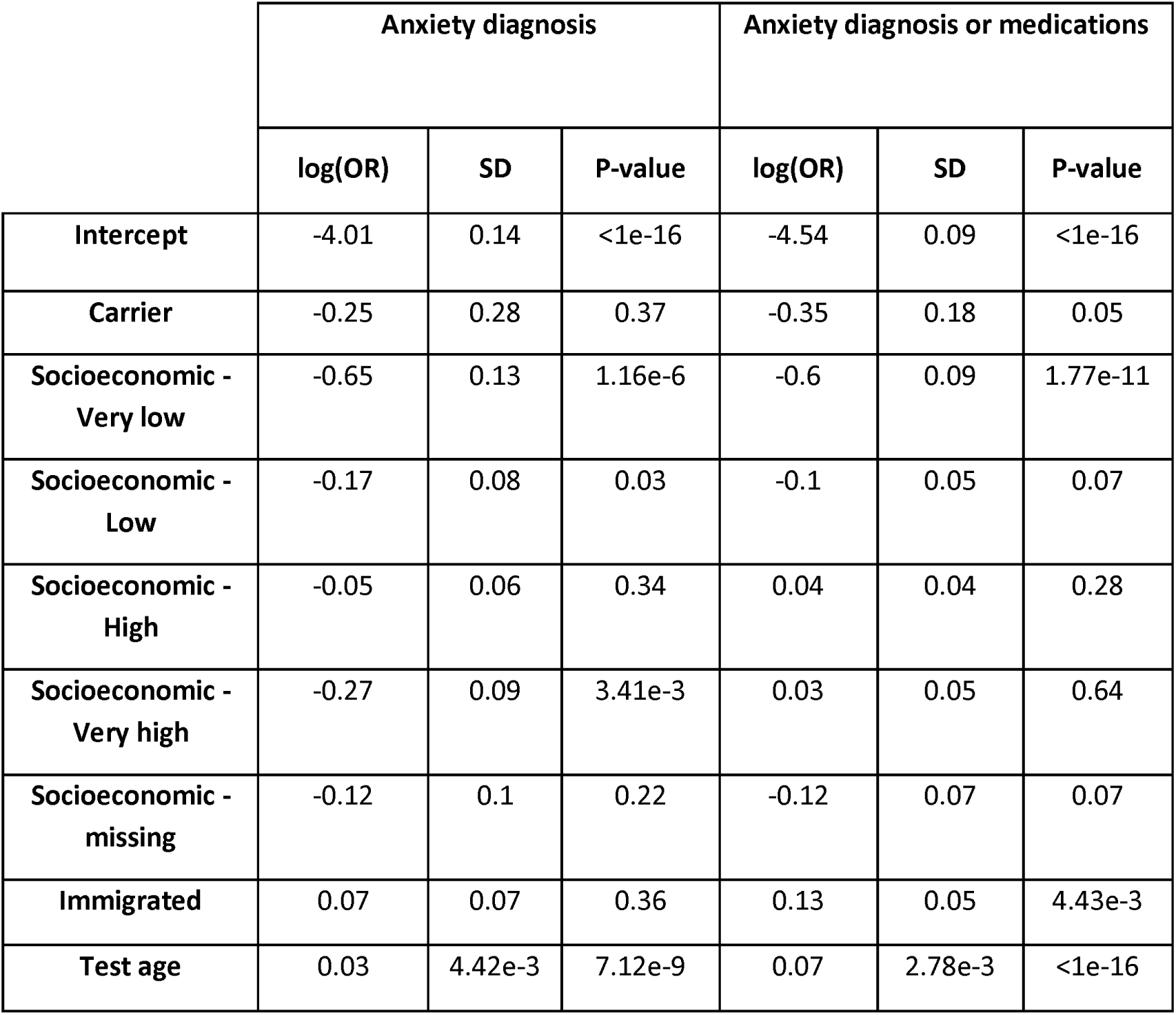
Logistic regression for anxiety.

## Notes

### Competing Interest Statement

The authors have declared no competing interest.

